# A Tale of Two Countries: Comparison of Rectal Cancer Characteristics Between Pakistani Americans and Native Pakistanis

**DOI:** 10.64898/2026.04.07.26350364

**Authors:** Maryam Sherwani, Mohammad Kumael Azhar, Sadaf Khan, Danish Ali, Syed Husain, Aimal Khan

## Abstract

**Introduction:** Comparison of rectal cancer characteristics in Pakistani Americans and native Pakistanis remains poorly investigated, as migrant studies have predominantly concentrated on East and Southeast Asian groups. This research aims to compare clinicopathological characteristics between the two groups. We hypothesize that significant differences will exist between these cohorts, mediated by gene-environment interactions.

**Methods:** This was a retrospective cohort study utilizing two multi-institutional databases to identify adult patients with rectal cancer: the National Cancer Database in the U.S (2018–2022) and the Rectal Cancer Surgery and Epidemiology Study in Pakistan (2020–2021). Non-Hispanic Whites (NHWs) were included as a reference population for comparative analysis. Clinicopathological characteristics were compared using Wilcoxon rank-sum and chi-square tests.

**Results:** A total of 523 Pakistani Americans and 608 native Pakistanis were included in the study. The median age at diagnosis was 57 years in Pakistani Americans (IQR 48-68), 42 years (IQR 33-54) in native Pakistanis and 63 years in NHWs (IQR 54-73) (p < 0.001). Native Pakistanis presented with early-stage disease less often than Pakistani Americans and NHWs (5.3%, 25.1%, and 20.5%, respectively; p < 0.001) and had markedly higher rates of signet cell carcinoma (20.1%, 0.6%, and 0.4%, respectively; p < 0.001) and poorly differentiated tumors (29.0%, 10.4%, and 11.4%, respectively; p < 0.001).

**Conclusions:** This study found that Native Pakistanis with rectal cancer presented at a younger age and with more aggressive tumor characteristics compared to both Pakistani Americans and NHWs. Notably, Pakistani Americans displayed a distinct clinical profile, intermediate between both groups.

## Introduction

Rectal cancer varies in incidence and presentation across ethnic and geographic populations, reflecting the interplay of genetic, epigenetic, environmental, and lifestyle factors (1,2). Pakistani Americans represent a large and rapidly growing demographic in the US, yet there is limited understanding of how the interaction between their genetic background, and the Western environment shapes the clinical presentation of rectal cancer in this group. Comparing rectal cancer characteristics between Pakistani Americans and native Pakistanis offers a unique opportunity to evaluate environmental influences on the development of this malignancy. This study aims to compare clinicopathologic characteristics between these populations.

## Methods

This retrospective comparative cohort study used data from the National Cancer Database (NCDB) in the US (2018–2022) and the Rectal Cancer Surgery and Epidemiology Study (ReCESS) in Pakistan (2020–2021). The NCDB is a joint project of the Commission on Cancer (CoC), the American College of Surgeons, and the American Cancer Society. It captures >70% of cancer cases diagnosed annually in the U.S(3). ReCESS is a multi-institutional collaborative of fourteen tertiary (public and private) hospitals in Pakistan formed by the Pakistan Rectal Cancer Working Group (PRCWG). Data from both databases were accessed on 1^st^ August 2025 by the research team.

Due to the retrospective deidentified nature of the databases, Institutional Review Board approval and patient informed consent were not required. This study adhered to the Strengthening the Reporting of Observational Studies in Epidemiology (STROBE) reporting guideline (4) (Appendix).

From the NCDB, adults (≥18 years) of ‘Pakistani’ or ‘Asian Indian or Pakistani’ race diagnosed with rectal adenocarcinoma were included as Pakistani Americans. Non-Hispanic Whites (NHWs) were included for reference. From ReCESS, adults with rectal cancer were included as Native Pakistanis.

Variables included age, sex, tumor histology, tumor grade (I-IV), clinical stage (American Joint Committee on Cancer 8th edition), and lymphovascular and perineural invasion. As continuous variables were non-normally distributed, they were summarized as median (interquartile range, IQR) and compared between groups using the Wilcoxon rank-sum test Categorical data were presented as frequencies and compared using the Chi-square test. Groups were compared using Wilcoxon rank-sum tests, with significance at p<0.05.

## Results

Between 2018 and 2022, 127,692 patients were identified with rectal cancer in the NCDB; after exclusions 523 Pakistani Americans and 92,366 NHWs remained. ReCESS included 613 patients with rectal cancer; 5 were excluded for being <18 years, resulting in a final sample of 608 Native Pakistanis. Missingness for each variable is provided in Supplementary Material (Table S1).

Median age differed significantly: 42 years for Native Pakistanis, 57 years for Pakistani Americans, and 63 years for NHWs. Native Pakistanis more frequently presented with advanced-stage disease (87.8%) than Pakistani Americans (57.2%), and NHWs (59.5%). Native Pakistanis had higher rates of signet-ring cell carcinoma (20.1%, 0.6%, and 0.4%) and poorly differentiated tumors (29.0%, 10.4%, and 11.4%) than Pakistani Americans and NHWs, respectively (Table 1 and Figure 1). In contrast, sex distribution, lymphovascular invasion, and perineural invasion rates did not differ significantly among the three groups.

**Table 1:** Comparison of demographic and clinicopathological characteristics between the study cohorts.

| Variables | Cohorts |  |  | p value |  |  |
| --- | --- | --- | --- | --- | --- | --- |
|  | Native Pakistanis (n=608) | Pakistani Americans (n=523) | NHWs (n=92,366) | Native vs Pakistani Americans | Native Pakistanis vs NHWs | Pakistani Americans vs NHWs |
| Age, median (IQR), years | 42 (21) | 57 (20) | 63 (19) | **<0.001** | **<0.001** | **<0.001** |
| Sex, n (%) |  |  |  |  |  |  |
| Female | 223 (36.9) | 201 (38.4) | 37,304 (40.4) | 0.601 | 0.083 | 0.364 |
| Clinical stage, n (%) |  |  |  |  |  |  |
| 0 | - | 7 (1.7) | 2,154 (3.0) | **<0.001** | **<0.001** | **0.016** |
| 1 | 25 (5.3) | 98 (23.4) | 12,564 (17.5) |  |  |  |
| 2 | 32 (6.8) | 74 (17.7) | 14,264 (19.9) |  |  |  |
| 3 | 352 (75.2) | 152 (36.4) | 26,953 (37.6) |  |  |  |
| 4 | 59 (12.6) | 87 (20.8) | 15,683 (21.9) |  |  |  |
| Histology, n (%) |  |  |  |  |  |  |
| Adenocarcinoma | 456 (75.0) | 429 (82.0) | 79,601 (86.2) | **<0.001** | **<0.001** | **0.023** |
| Signet-ring cell | 122 (20.1) | 3 (0.6) | 371 (0.4) |  |  |  |
| Others | 30 (4.9) | 91 (17.4) | 12,394 (13.4) |  |  |  |
| Grade, n (%) |  |  |  |  |  |  |
| Well differentiated | 65 (12.2) | 55 (15.0) | 7,754 (13.8) | **<0.001** | **<0.001** | 0.715 |
| Moderately differentiated | 313 (58.8) | 273 (74.6) | 42,112 (74.7) |  |  |  |
| Poorly differentiated | 154 (29.0) | 38 (10.4) | 6,399 (11.4) |  |  |  |
| Undifferentiated | - | 0 (0.0) | 97 (0.2) |  |  |  |
| Lymphovascular invasion, n (%) | 45 (15.1) | 44 (19.0) | 8,251 (19.1) | 0.231 | 0.074 | 0.949 |
| Perineural invasion, n (%) | 38 (12.7) | 36 (15.5) | 6,468 (15.0) | 0.365 | 0.261 | 0.861 |

Bold p-values are significant at a 0.05 level.

**Figure 1:**
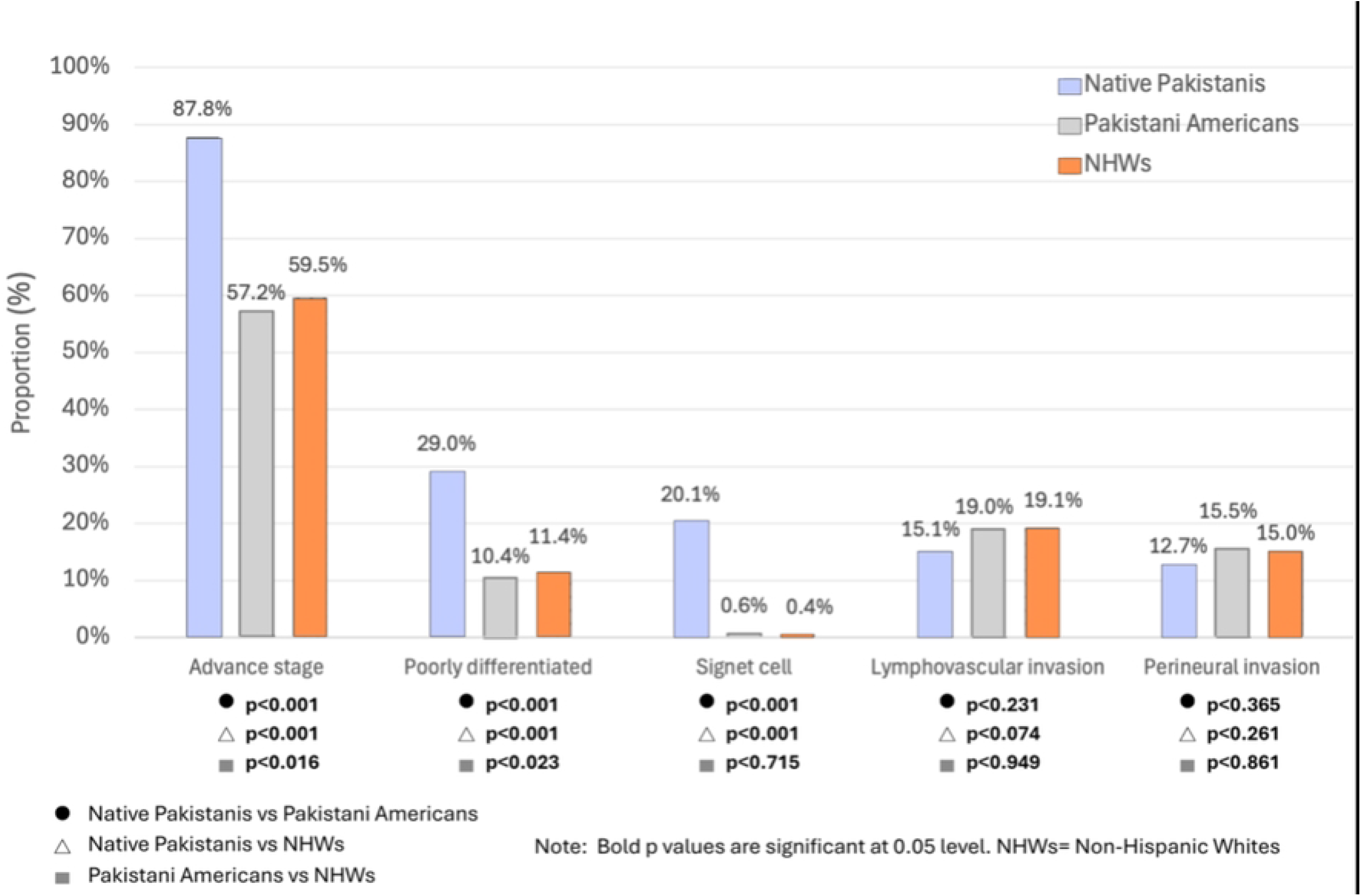
Comparison of rectal cancer characteristics among Native Pakistanis, Pakistani Americans and NHWs

## Discussion

Native Pakistanis with rectal cancer presented at a younger age and with more aggressive tumor characteristics than Pakistani Americans and NHWs. Pakistani Americans exhibit intermediate disease characteristics, suggesting that migration and acculturation influence disease presentation.

Signet-ring cell carcinoma was strikingly more frequent in native Pakistanis (20.1%) compared with NHWs and Pakistani Americans (<1%). This histology is associated with poor prognosis and association with younger age, which aligns with our findings(5,6). Notably, the median age at diagnosis in this group (42 years) falls below current screening thresholds, supporting reconsideration of screening strategies in high-risk populations.

The aggressive phenotype observed in Native Pakistanis was attenuated in Pakistani Americans. Earlier stage at diagnosis may reflect enhanced access to colorectal cancer screening in the US; however, the shift in tumor histology and grade likely represents gene-environment interaction. While incidence may also be increasing, this could not be estimated. It is plausible that Western environmental influences, while contributing to an increased overall risk, may concurrently favor less aggressive tumor phenotypes.

This study has several limitations. The use of the combined “Asian Indian or Pakistani” race category in NCDB may have introduced misclassification bias. Similarly, in the ReCESS database, all patients were assumed to be native Pakistanis. Data accuracy depended on the reporting quality of the databases, and coding errors, missing data, or inconsistencies may have impacted results.

Rectal cancer varies even among populations of shared ethnic origin, underscoring the effects of epigenetic modifications. These findings have implications for both developed nations, where screening is available yet underutilized among minority groups, and developing countries, where accelerating dietary Westernization, concurrent with limited screening infrastructure may foreshadow a growing burden of early-onset and unpredictable disease(7,8).

## Data Availability

The data that support the findings of this study are available on request from the corresponding author, AK. The data are not publicly available due to license restrictions of the NCDB and ReCESS databases.

## Acknowledgements

None

